# SARS-CoV-2 new infections among health care workers after the first dose of the BNT162b2 mRNA Covid-19 Vaccine

**DOI:** 10.1101/2021.03.24.21254238

**Authors:** Carlos Guijarro, Isabel Galán, Diana MartÍnez-Ponce, Elia Pérez-Fernández, Maria José Goyanes, Virgilio Castilla, MarÍa Velasco

## Abstract

**Objectives:** To evaluate the effect of mRNA SARS-Cov-2 vaccination on the incidence of new SARS-CoV-2 infections in health care workers (HCW).

**Methods:** The evolution of the incident rate of microbiologically confirmed SARS-CoV-2 infection in a cohort of 2590 HCW after **BNT162b2** mRNA SARS-CoV-2 vaccination, as compared to the rate in the community (n=170513) was evaluated by mixed Poisson regression models

**Results:** A total of 1820 HCW (70,3% of total) received the first dose of the **BNT162b2 mRNA** vaccine between January 10-16, 2021), and 296 (11,4%) the following week. All of them completed vaccination 3 weeks later. Incidence rates of SARS-CoV-2 infection after the first dose of mRNA SARS-CoV-2 vaccine declined by 71% (Incidence Rate Ratio (IRR) 0.286, 95% confidence interval (CI) 0.174-0.468, p< 0.001) and by 97% (IRR 0.03 95% CI 0.013-0.068, p<0.001) after the second dose as compared to the perivaccine time. SARS-CoV-2 incidence rates in the community (with a negligible vaccination rate) had a much lower decline: 2% (IRR 0.984; 95% CI 0.943-1.028; p = 0.47) and 61% (IRR 0.390, 95% CI 0.375-0.406; p<.001) for equivalent periods. Adjusting for the decline in the community, the reduction in the incident rates among HCW were 73% (IRR 0.272; 95% CI 0.164-0.451 p<0.001) after the first dose of the vaccine and 92 % (IRR 0.176, 95% CI 0.033-0.174; p<0.001) after the second dose.

**Conclusions:** mRNA SARS-CoV-2 vaccination is associated with a dramatic decline in new SARS-CoV-2 infection among HCW, even before the administration of the second dose of the vaccine.

## Introduction

Besides the appropriate use of personal protection equipment, vaccination has become the main tool for the control of SARS-COV-2 infection in the clinical setting [1]. However, there is scant evidence regarding the effects of SARS-CoV-2 vaccination on the incidence of new SARS-CoV-2 infection among health care workers (HCW) [2–5].

We sought to describe the effects of a hospital wide vaccination program with**BNT162b2** mRNA COVID-19 vaccine on the evolution of new SARS-CoV-2 infections among HCW, as compared to the evolution in the general population in the same geographical area.

## Methods

Cohort of 2590 HCW attended at the Occupation Health Unit of the hospital. SARS-CoV-2 tests from nasopharyngeal swabs were performed for all HCW when they had a SARS-CoV-2 risk exposure (either at work or in the community) or developed any symptoms compatible with SARS-CoV-2 infection. Abbott Panbio Rapid Antigenic Corona® antigen test was used for symptomatic HCW[6]. If negative, a SARS-CoV-2 PCR test was performed 48-72 hours thereafter. Asymptomatic HCW were evaluated by SARS-CoV-2 PCR test in nasopharyngeal swabs with Bio-Rad CFX96™ Real-Time PCR Detection System, as previously described [7,8]. New HCW infections were defined as a SARS-CoV-2 positive antigen or PCR test. Date of infection was defined as the date of the first positive test. Weekly incident rates and 95% exact Poisson confidence intervals (CI) were calculated. Community weekly incidence rates at Alcorcón (n= 170513 inhabitants) were obtained from the official data published by regional and national authorities[9]. For the purpose of this paper, three time periods were used: peri-vaccination time (Period 0: three weeks before the beginning of vaccination, the first week of first dose massive vaccination, and the week thereafter), intermediate (Period 1: weeks 2-4 after 1st dose vaccination), and post-vaccination, (Period 3: weeks 5-14 after the first doses of the vaccine, i.e. 1 week after the full vaccination and thereafter; Figures 1 & 2). These times were chosen as representative of baseline pre-vaccination, and expected immunological effects of the first dose of the vaccine and the full vaccination [10]. Relative changes in the incidence of new SARS-CoV-2 infection in HCW and in the geographical area of the Hospital were evaluated by multivariate regression models. To estimate rate ratio between periods, a mixed Poisson regression model with group (HCW/community) and time periods as random effect and group population as exposure variable was adjusted. Analysis was performed with STATA 14. The study was approved by the Independent Review Board (CEIm) of Hospital Universitario Fundación Alcorcón All HCW signed an informed content.

**Figure 1.**
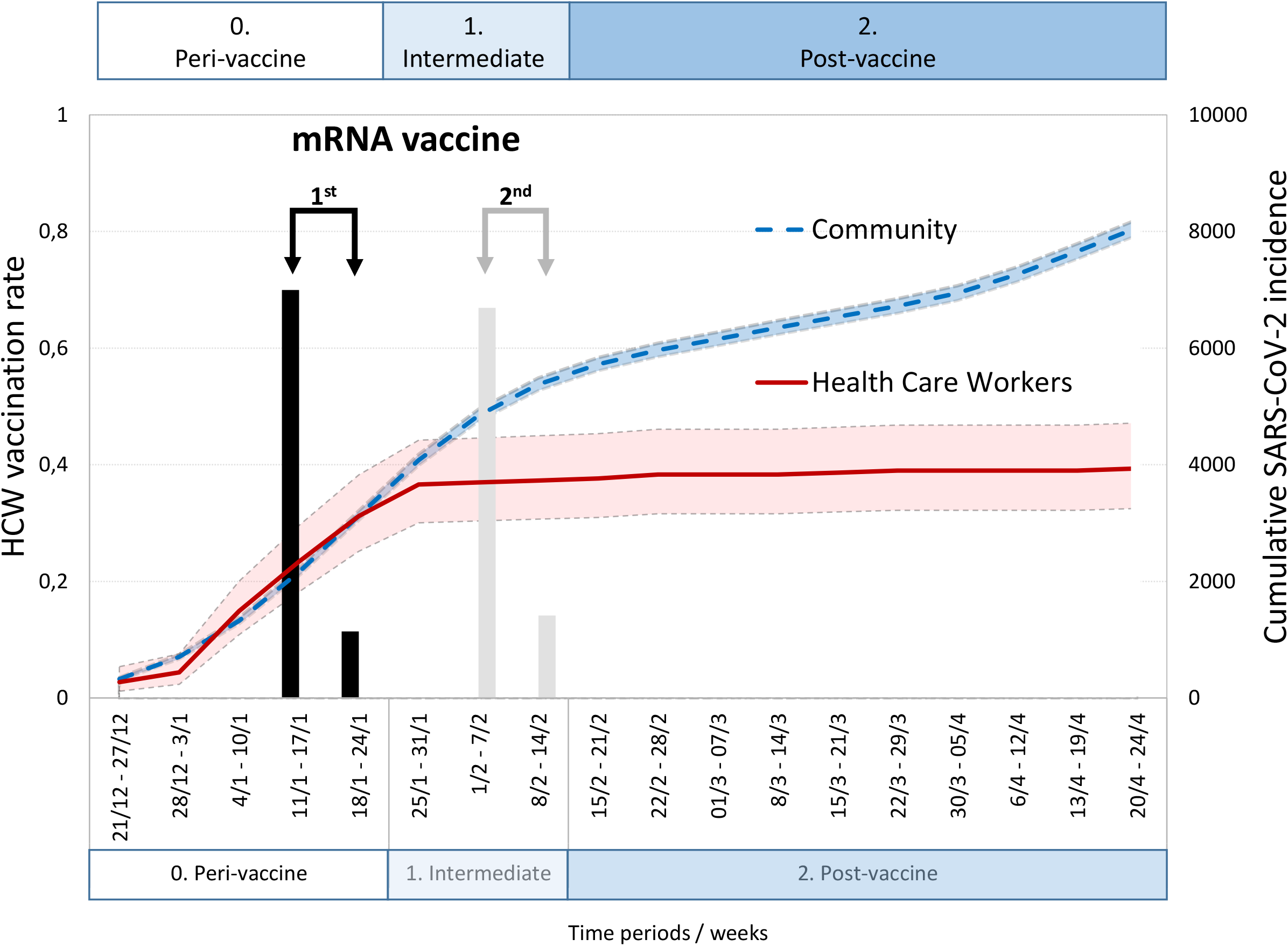
Cumulative incident rate of new SARS-CoV-2 infections among Health Care Workers (HCW) at Hospital Universitario Fundación Alcorcón and the general population at Alcorcón. Cumulative incident rate per 100000 individuals of new SARS-CoV-2 infections among Health Care Workers (HCW) at Hospital Universitario Fundación Alcorcón (n=2590 ⍰⍰) and the general population at Alcorcón (Madrid, Spain; n=170513 ‐) from December 21, 2020. Shaded areas depict 95% confidence intervals. The proportion of HCW receiving the first and second dose of the BNT162b2 mRNA Covid-19 Vaccine are indicated at the appropriate times (bars). Vaccine-related periods are highlighted indicating the expected immunological effects of the vaccine.

**Figure 2.**
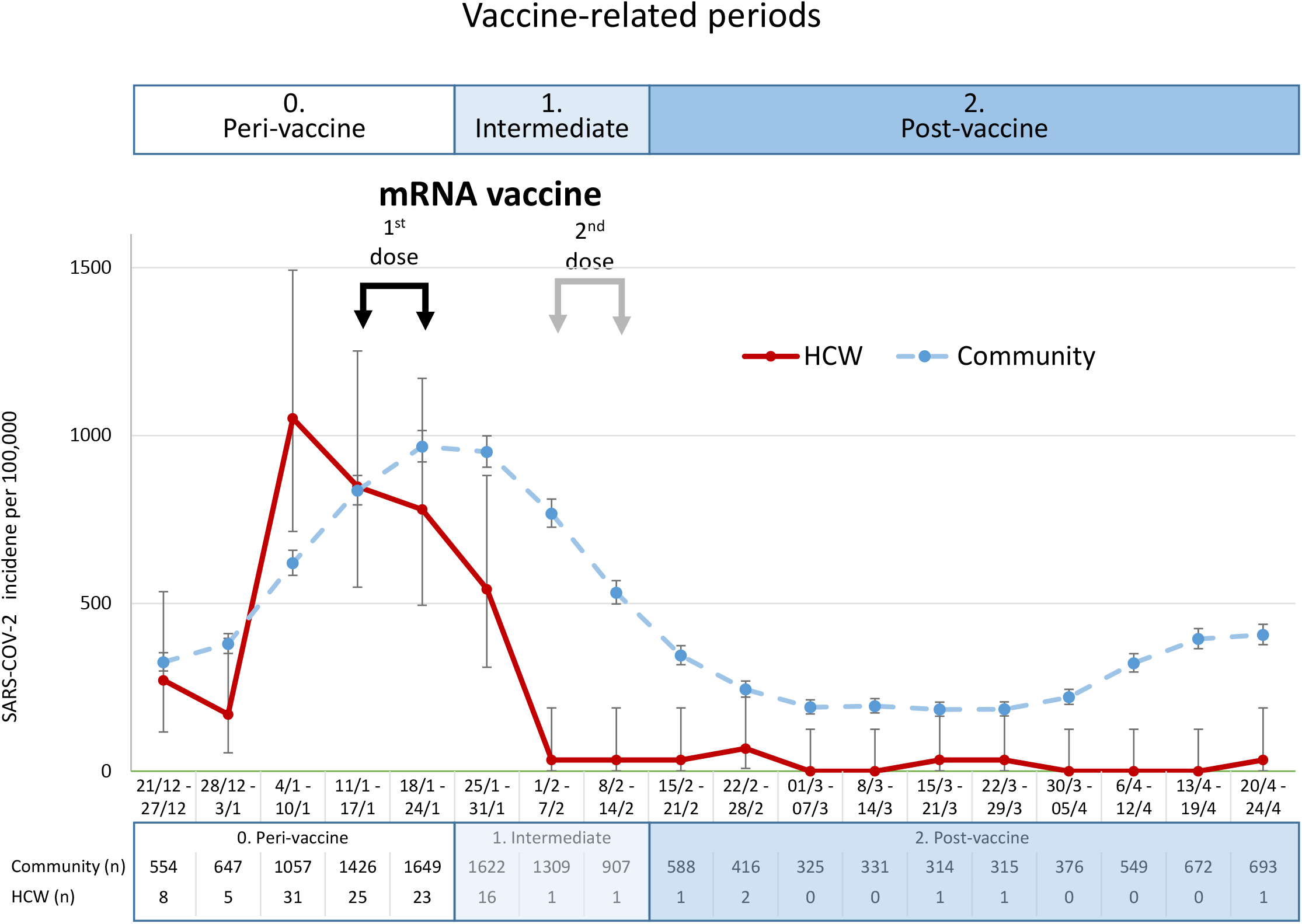
Legend. Weekly incident rate of new SARS-CoV-2 infections among Health Care Workers (HCW) at Hospital Universitario Fundación Alcorcón and the general population of Alcorcón. Weekly incident rate per 100000 individuals of new SARS-CoV-2 infections among Health Care Workers (HCW) at Hospital Universitario Fundación Alcorcón (n=2590) and 95% exact Poisson confidence intervals. As a reference, the incident rate and the general population at Alcorcón (Madrid, Spain; n=170513 ---) depicts the evolution of the ‘third’ wave of the disease. Bottom table describes the weekly individual new diagnosis (n) of SARS-CoV2-infections both among HVW and the general population of Alcorcón. Timing of the first and second dose of the BNT162b2 mRNA Covid-19 Vaccine are indicated (arrows). Vaccine-related periods are highlighted indicating the expected immunological effects of the vaccine.

## Results

The BNT162b2 COVID-19 mRNA vaccine was offered to all HCW as soon as it was available (January 11, 2021). A total of 1820 HCW (70% of total) received the first dose in the first week (11-17 January 2021), and 296 (11%) the following week (Figure). A total of 116 new SARS-CoV infections were detected at the Occupational Health Unit among HCW during the study period (between December 21^st^ 2020, and April 24 2021. Seventy one HCW (61%) had a positive PCR test, 18 (16%) a positive antigen test, and 27 (23%) both. Globally, 84% of new infections were confirmed by at least one positive PCR test. Fifty five new infections were symptomatic (47%), while the remaining 60 (53%) were detected as a part of a screening protocol after close contact with COVID-19 cases (n=29; 25%), nosocomial outbreak evaluation (n=29; 25%) or travel requirements (n=2; 2%).

The number of HCW diagnosed of acute SARS-CoV-2 infection dropped by 71% in the weeks 2-4 after first dose vaccination as compared to the previous 5 weeks (Period 0. Perivaccine time), Incidence Rate Ratio (IRR) 0.286, 95% confidence interval (CI) 0.174-0.468, p< 0.001; Figures 1 &2:; ; Supplemental Table and Figure). New SARS-CoV-2 infections among HCW virtually disappeared after the second dose dose of the vaccine (97% reduction, IRR 0.030 95% CI 0.013-0.068, p<0.001) as compared to the Period 0 (perivaccine time; Figures 1 &2; Supplementary Table 1). There was also a reduction of new SARS-COV-2 infections in the general population at Alcorcón (Figures 1 & 2), depicting the evolution of the &‘third’ wave of the disease 2% (IRR 0.984; 95% CI 0.943-1.028; p = 0.47) and 61% (IRR 0.390, 95% CI 0.375-0.406; p<.001) for the same time periods. During the time of the study the vaccination rate in the community was negligible (4,8% on March 29 [11]). By Poisson model regression analysis, the reduced incidence among HCW adjusting for the background changes in the general population remained statistically and clinically significant: 3% reduction (IRR 0.272; 95% CI 0.164-0.451 p<0.001) after the first dose of the vaccine and 92 % (IRR 0.176, 95% CI 0.033-0.174; p<0.001) after the second dose (Supplemental Tables and Figure). It is conceivable that the response to the vaccine among HCW may have been boosted by a previous exposure to the virus [12,13].. As a sensitive analysis we restricted the evaluation of new SARS.CoV-2 infections to HCW (n=1582) who had never tested positive in any of our wide seroprevalence surveys [7]. As shown in the supplementary table 2, the decline in new SARS-CoV-2 infections was essentially identical to the total HCW population.

## Discussion

Our results show a dramatic reduction in new SARS-CoV-2 infections in HCW after a hospital-wide vaccination program with the BNT162b2 mRNA Covid-19 vaccine. Interestingly, the decline was important even before the administration of the second dose of the vaccine, suggesting that a single dose may confer a substantial proportion, in agreement with the original mRNA vaccination trials as well as recent observational studies [2–5]. However, our results should be interpreted with caution. First, we report a limited follow-up in a medium size hospital. Second, there was also a significant decline in the SARS-CoV-2 incidence in the general population in this time frame. The important reduction in the community rate most likely reflects general public health measures by regional and national governments, since the rate of vaccination in the general population in (Madrid) was lower than 5% [11]: Therefore, its potential effect on the declining infection rates in the community should be marginal. The powerful reduction among HCW strongly suggests a main role for the vaccine.

Our results after a single dose of the vaccine might not be extrapolated to other settings: our vaccination program was extended to all HCW, including HCW with previous SARS-COV-2 documented infection [7]. We and others have recently shown that HCW with a previous SARS-CoV-2 infection exhibit a strong serologic response to the first dose of the vaccine, reaching higher IgG anti spike titre than that obtained after full vaccination in SARS-CoV-2 naïve individuals [12,13]. Other vaccination programs (excluding previously infected HCW) may exhibit a lower protection [2]. The protective effect of the first dose of the vaccine in settings with a lower seroprevalence such as the general population in Madrid may be less evident [14]. However, in a sensitive analysis by excluding SARS-CoV-2 seropositive HCW before vaccination, the estimated decline in SARS-CoV-2 infection remained similar to the total HCW population. Recent reports have described a high degree of protection in HCW receiving the first dose of an RNA Vaccine, in agreement with our results [5]. Our results are restricted to the BNT162b2 mRNA vaccine. Whether similar outcomes may be obtained with other vaccines cannot be ascertained from our data. However, recent data from the UK suggest that the first dose of mRNA or other vaccines may be substantial [15]. The optimal program vaccination for HCW with previous SARS-CoV-2 infection remains to be defined.

In conclusion, a wide vaccination program for HCW in the real world seems to offer a powerful protection from new SARS-CoV-2 infection and provides a much safer clinical environment. Significant declines in SARS-CoV-2 infection rates may be achieved even before the full vaccination program is completed. These data may encourage hesitant HCW to join vaccination efforts [4].

## Data Availability

Data may be available upon request

## Acknowledgments

The authors would like to acknowledge the work of nurses, administrative personnel, pharmacists and doctors that made possible the deployment of the vaccination program.

## Contribution

Writing - Original Draft: CG and MV.; Review & Editing: CG, IG, DMP, EPF, JG, ZC, MVA; Conceptualization: CG, IG, VC and MV; Data acquisition: IG, DMP, MJG. EPF Methodology: CG, EPF, MJG, MV.; Formal Analysis: CG, EPF, and MV;

---

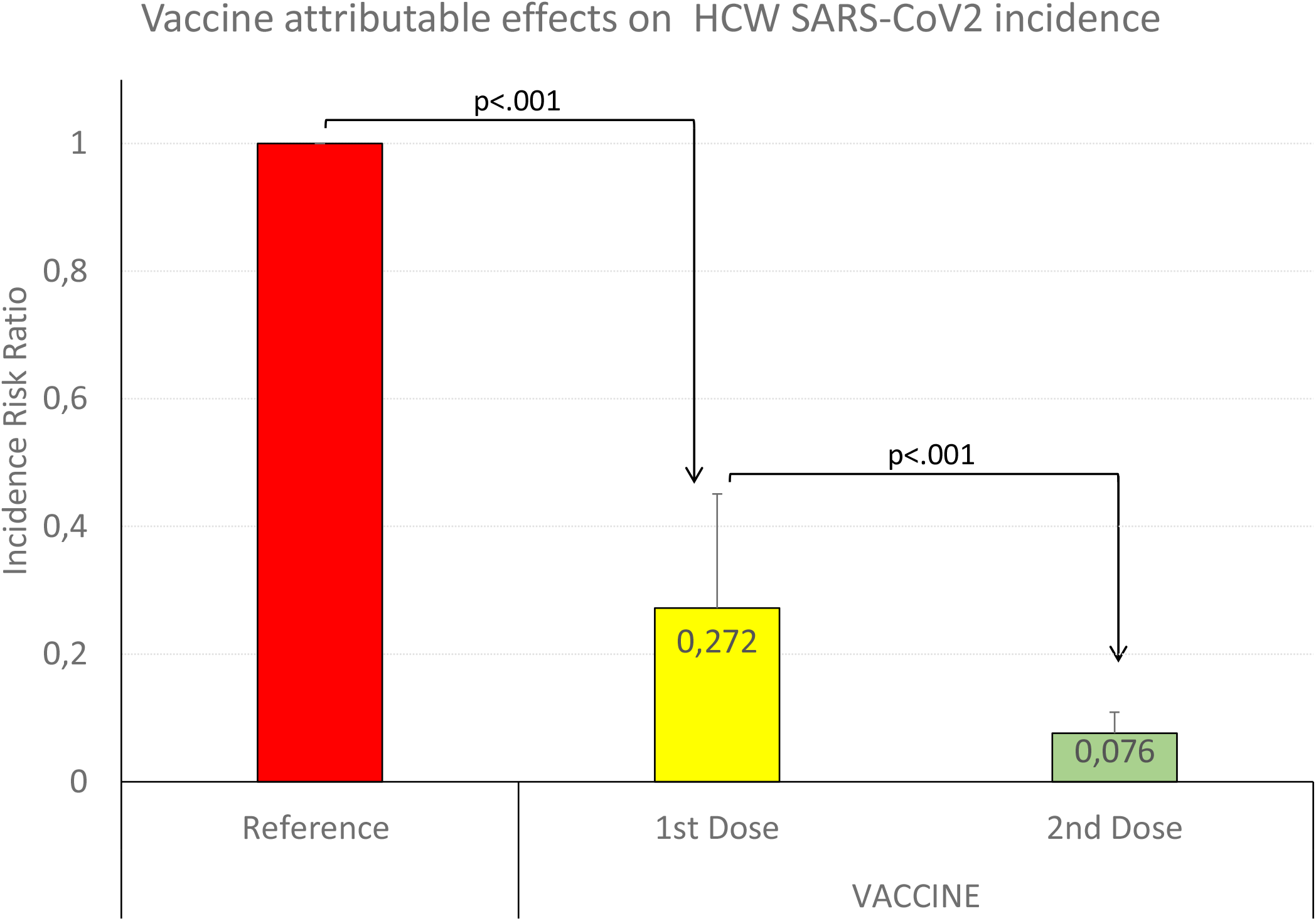

