## Supplement for "SARS-CoV-2 new infections among health care workers after the first dose of the BNT162b2 mRNA Covid-19 Vaccine"

Supplementary Table 1. Changes in the incidence rate ratio of SARS-CoV-2 infection among HCW and the community

| | | **A. IRR by period and population group** | | | | | **B. IRR by period in HCW attributable to the vaccine** | | | | |
|---|---|---|---|---|---|---|---|---|---|---|---|
| **Population** | **Period** | **IRR** | **p-value** | **95 % CI%** | | **Relative change** | **IRR** | **p-value** | **95 CI%** | | **Relative change** |
| **COMMUNITY** | **Intermediate** | 0.984 | 0.474 | 0.943 | 1.028 | -1.6% | | | | | |
| | **Post-vaccine** | 0.390 | <0.001 | 0.375 | 0.406 | -61.0% | | | | | |
| **HCW** | **Intermediate** | 0.268 | <0.001 | 0.161 | 0.444 | -73.2% | 0.272 | <0.001 | 0.164 | 0.451 | -72.8% |
| | **Post-vaccine** | 0.030 | <0.001 | 0.013 | 0.068 | -97.0% | 0.076 | <0.001 | 0.033 | 0.174 | -92.4% |
| IRR: Incidence rate ratio; HCW: Heatlh care worker; CI confidence interval | | | | | | | | | | | |

Mixed Poisson regression model with group (HCW/community) and time periods as random effect and group population as exposure variable was adjusted to estimate the rate ratio for SARS-CoV-2 infection among HCW and the community for vaccine related periods. (A). Time periods were chosen as described in main text and figure to evaluate the expected effect of the first and the second dose of the BNT162b2 mRNA Covid-19 Vaccine among HCW. B, Incidence Rate ratio for HCW after adjusting for incidence changes in the community. The interaction effect estimated with Mixed Poisson model was 0.27 (CI95% 0.164-0.451, p<0.001) in intermediate period and 0.076 (CI95%: 0.033-0.174, p<0.0001) in post-vaccine period. This effect can be interpreted as change from peri-vaccine period in HCW attributable to the vaccine, with a reduction of 72.8% and 92.4% in intermediate and post-vaccine periods respectively (Supplemental Figure) .

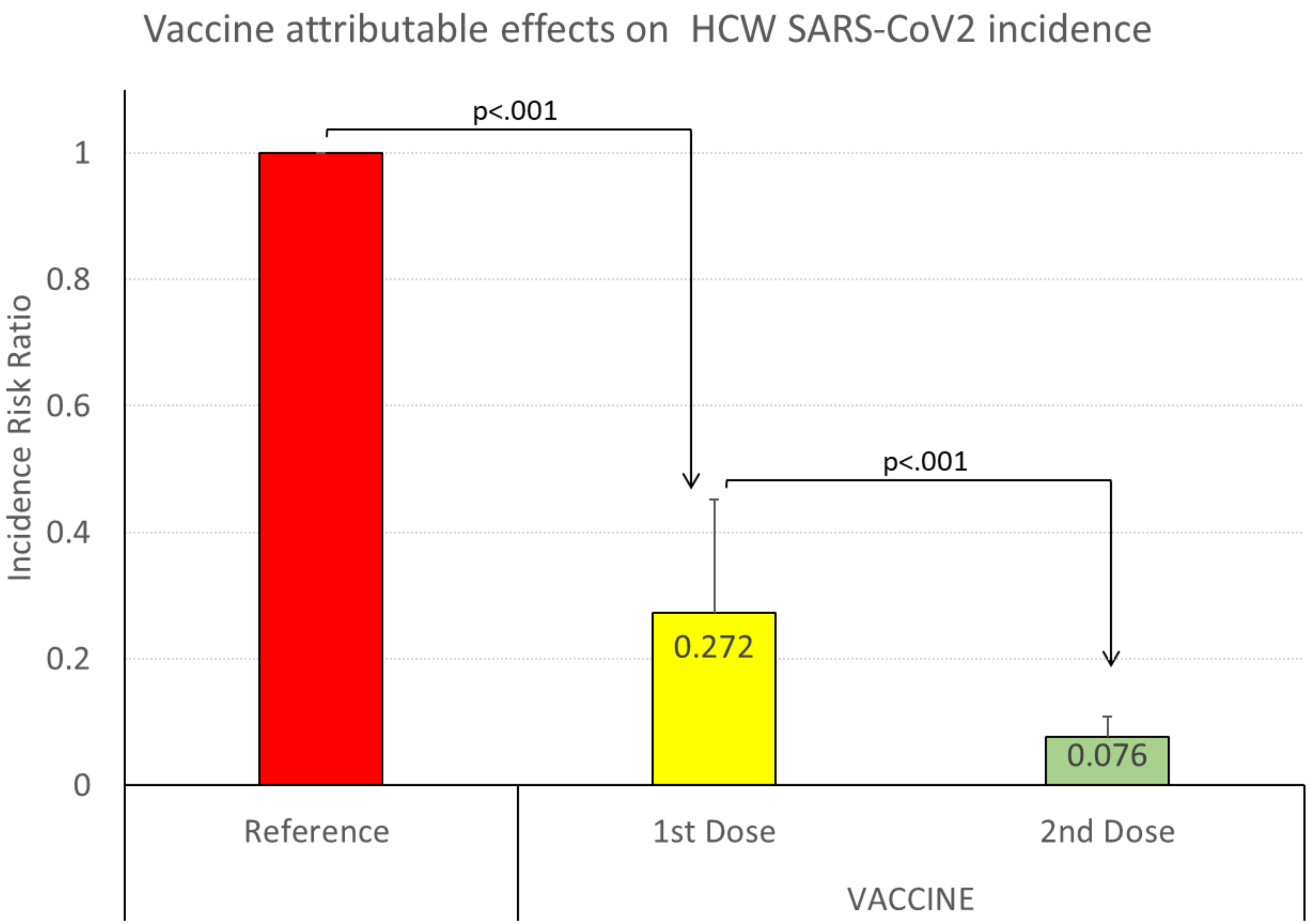


See table legend for details

Supplementary Table 2. Sensitive Analysis.

| Population | Period | IRR by period and population group. Sensitive analysis | | | | |
|---|---|---|---|---|---|---|
| | | IRR | p-value | 95% CI | | Relative change |
| **COMMUNITY** | **Intermediate** | 0.985 | 0.486 | 0.944 | 1.028 | -1.5% |
| | **Post-vaccine** | 0.390 | <0.001 | 0.375 | 0.406 | -61.0% |
| **HCW** | **Intermediate** | 0.258 | <0.001 | 0.143 | 0.466 | -74.2% |
| | **Post-vaccine** | 0.033 | <0.001 | 0.013 | 0.082 | -96.7% |
| iRR: Incidence rate ratio; HCW: Health Care Worker; CI: Confidence Interval | | | | | | |

Mixed Poisson regression model with group (HCW/community) and time periods as random effect and group population as exposure variable was adjusted to estimate the rate ratio for SARS-CoV-2 infection among HCW and the community for vaccine related periods. For this analysis HCW with a positive serologic test for SARS-CoV-2 in any of the two hospital wide seroprevalence studies were excluded (n=1582). Time periods were chosen as described in main text and figures to evaluate the expected effect of the first and the second dose of the BNT162b2 mRNA Covid-19 Vaccine among HCW. Results are essentially identical as those reported for the full HCW population (Table 1).
